# Development of Prediction Models of COVID-19 Vaccine Uptake among Lebanese and Syrians in a district of Beirut, Lebanon: a population-based study

**DOI:** 10.1101/2024.03.21.24304659

**Authors:** Marie-Elizabeth Ragi, Hala Ghattas, Hazar Shamas, Jocelyn DeJong, Nada M. Melhem, Stephen J. McCall, with the CAEP Study Group

## Abstract

**Introduction:** Vaccines are essential to prevent infection and reduce morbidity of infectious diseases. Previous evidence has shown that migrants and refugees are particularly vulnerable to exclusion and discrimination, and low COVID-19 vaccine intention and uptake were observed among refugees globally. This study aimed to develop and internally validate prediction models of COVID-19 vaccine uptake by nationality.

**Methods:** This is a nested prognostic population-based cross-sectional analysis. Data was collected between June and October 2022 in Sin-El-Fil, a district of Beirut, Lebanon. All Syrian adults and a random sample of adults from low-socioeconomic status neighborhoods were invited to participate in a telephone survey. The main outcome was uptake of COVID-19 vaccine. Predictors of COVID-19 vaccine uptake were assessed using LASSO regression for Lebanese and Syrian nationalities, respectively.

**Results:** Of 2,045 participants, 79% were Lebanese, 18% Syrians and 3% of other nationalities. COVID-19 vaccination uptake was higher among Lebanese (85% (95%CI:82-86) compared to Syrians (47% (95% CI:43-51)) (P<0.001); adjusted odds ratio (aOR) 6.8 (95%CI:5.5-8.4). Predictors of uptake of one or more vaccine dose for Lebanese were older age, presence of an older adult in the household, higher education, greater asset-based wealth index, private healthcare coverage, feeling susceptible to COVID-19, belief in the safety and efficacy of vaccines and previous receipt of flu vaccine. For Syrians they were older age, male, completing school or higher education, receipt of cash assistance, presence of comorbidities, belief in the safety and efficacy of vaccines, previous receipt of flu vaccine, and legal residency status in Lebanon.

**Conclusions:** These findings indicate barriers for vaccine uptake in Syrian migrants and refugees, including legal residency status. They call for urgent action to enable equitable access to vaccines by raising awareness about the importance of vaccination and the targeting of migrant and refugee populations through vaccination campaigns.

**Key Messages:** *What is already known on this topic:* Vaccines are essential to prevent infection and reduce morbidity of infectious diseases, and vulnerable populations may lack access to vaccination campaigns.

*What this study adds:* To the best of our knowledge, no studies have examined compared predictors of COVID-19 vaccine uptake and measured the rate of vaccination among between Syrian migrants and refugees and their Lebanese host communities. This study illustrates a clear difference in vaccine uptake between nationalities and developed prediction models among the Syrian and Lebanese that identified differential predictors of COVID-19 vaccine uptake for each population.

*How this study might affect research, practice or policy:* These findings indicate barriers for vaccine uptake in Syrian migrants and refugees, including legal status, and calls for urgent action to enable access to vaccines by raising awareness about the importance of vaccination against COVID-19 in vulnerable groups and targeting migrant and refugee populations through vaccination campaigns.

## Introduction

Vaccines were an essential public health tool to mitigate the impact of the COVID-19 pandemic. In Lebanon, vaccines were available free of charge to all residents starting February 2021 following the WHO SAGE risk-and age-based approach for prioritization (1–4). The COVID-19 national vaccination plan aimed to reach community immunization levels of at least 70-80% by the end of 2022 (2–4), yet there were subpopulations that remained unvaccinated. A survey amongst older Syrian refugees in Lebanon showed that only 42.5% were vaccinated against COVID-19 by March 2022 (5), while the national average for all residents of Lebanon was 50% by October 2022 (6).

Lebanon hosts the largest number of refugees per capita in the world, including Syrian, Palestinian and Iraqi refugees, that account for approximately one third of the population, many of which are dispersed in urban residential areas living amidst Lebanese communities (4, 7). Migrants and refugees are particularly vulnerable to exclusion, stigma and discrimination that may limit access to vaccination campaigns (7). Required renewal of residency permits per year at significant economic costs adds additional burden (8). Studies in other settings have shown that refugees and non-native ethnic groups were less willing to get vaccinated against COVID-19 as compared to native populations (9–13), with racial discrimination and lack of trust in authorities’ facilities and systems being identified as determinants of COVID-19 vaccine hesitancy (12, 13). Thus, the factors that impact vaccine uptake are likely to differ between national and non-national populations, and understanding demographic variations and predictors of vaccination status between these subpopulations remains important to inform the response to future outbreaks of COVID-19 and future pandemics.

This study aims to determine whether there are differences in COVID-19 vaccine uptake by nationality in a suburb of Beirut, Lebanon, and to develop and internally validate prediction models that examine context-specific differential predictors of COVID-19 vaccine uptake by nationality.

## Methods

### Study design and setting

This is a nested cross-sectional analysis within an ongoing multi-wave longitudinal study that aims to track vulnerabilities to COVID-19 and other health emergencies in Sin-El-Fil, a suburb of Beirut, Lebanon. The parent study focused on subpopulations with vulnerabilities that may increase risk of COVID-19 infection, morbidity or mortality. These included 1) adults aged 60 years or older, 2) pregnant women, 3) adults living in areas of low socioeconomic status, and 4) Syrian migrants and refugees. The study protocol was reviewed and approved by the American University of Beirut Social and Behavioral Sciences Institutional Review Board (reference: SBS-2021-0268).

### Sampling and study population

This was a multi-stage stratified sampling design using an area-based sampling (14). The suburb of Sin-El-Fil was stratified into areas of low and high socioeconomic status, with boundaries defined geographically through stakeholder consultation with the Sin-El-Fil municipality and NGOs. A household listing exercise was carried out in April 2022 whereby all households in Sin-El-Fil were enumerated face-to-face to complete an eligibility screening survey, which identified households and individuals with the specific vulnerability criteria mentioned above (15). Pre-consent was obtained from all households with eligible individuals and phone numbers were collected from consenting households. For the study sample selection, a listing of all eligible household members was generated. A sample of older adults and a sample of adults between 18-60 years old were randomly selected using proportionate allocation. Several of the vulnerability groups were small so these group were oversampled; this included the selection of all Syrian adults and pregnant women. For the present analysis, the study population included all participants living in areas of low socioeconomic status who completed the first wave of data collection conducted between June and October 2022 (n=2,045).

Respondents were contacted to complete a computer assisted telephone survey conducted by a trained data collector and data were entered on SurveyCTO software (Dobility Inc., Cambridge, MA, USA). Verbal informed consent to participate in an oral telephone interview was obtained from selected respondents and those aged 60 years or older were furthermore assessed for capacity to participate using 5 modified items from the University of California, San Diego, Brief Assessment of Capacity to Consent (16).

### Data sources

The questionnaire was developed using existing validated scales and questionnaire modules, contextualized questions, and community-identified priorities. The survey was created in collaboration with representatives from the municipality of Sin-El-Fil, the Ministry of Public Health (MOPH), and Non-Governmental Organizations (NGOs) operating in Sin-El-Fil. The survey tool was drafted in English, translated to Arabic and tested for comprehension prior to deployment. Data were frequently monitored in parallel with data collection for quality assurance, with 5% call back checks conducted to ensure accuracy of the collected data.

### Outcome measures

COVID-19 vaccine uptake of at least one dose was the primary outcome of interest. Each participant was asked the following questions: “Have you received the COVID-19 vaccine?” and “If yes, how many doses did you receive?”. The outcome is presented as binary: “received the COVID-19 vaccine” and “did not receive the COVID-19 vaccine”. Additionally, unvaccinated participants were asked if they planned to take the COVID-19 vaccine and the reasons why not.

### Possible predictors

Based on the literature on COVID-19 vaccine acceptance and hesitancy, several possible predictors for vaccine uptake were included. For Lebanese, these were: age, sex, education, household assets wealth index, presence of an older adult in the household, presence of chronic illness, healthcare coverage, COVID-19 knowledge and risk perceptions (susceptibility to COVID-19), general perceptions of vaccine safety and effectiveness, and receipt of the flu vaccine. The household assets-based wealth index was generated through Principal Component Analysis (PCA) from variables on presence in the household of functional transportation, communication, home technology and cooking assets. The chronic illnesses assessed included hypertension, type II diabetes, vascular diseases, dyslipidemia, chronic respiratory diseases, rheumatoid arthritis, chronic kidney diseases, cancer. For Syrians, these were age, sex, education, receipt of cash assistance and legal residency status in the country that are context specific to Syrian migrants and refugees, presence of chronic illness, healthcare coverage, COVID-19 knowledge and risk perceptions, general perceptions of vaccine safety and effectiveness, and receipt of the flu vaccine. The missing values were assumed to be missing at random. The variable with the highest amount of missing values was at 6.3% and a complete case analysis was performed (17).

### Statistical analysis

Frequencies and percentages are presented for categorical variables, and medians with their interquartile range (IQR) for continuous variables. The analysis accounted for the complex design and non-response. Using unadjusted logistic regression analyses, the association between COVID-19 vaccine uptake and the possible predictors was examined among the different nationalities, unadjusted odds ratios (ORs) along with their 95% confidence intervals (CIs) are reported. The outcome was a binary variable for uptake of the COVID-19 vaccine. All variables were categorical except age, which was found to have a linear association with COVID-19 vaccine uptake.

A Least Absolute Shrinkage and Selection Operator (LASSO) logistic regression model was used to identify the predictors of vaccine uptake for each nationality (Lebanese and Syrian), and all corresponding candidate predictors were entered into each model. The final models were internally validated using cross-validation (10-folds). Multicollinearity was assessed using correlation matrices where a variance inflation factor (VIF) greater than five indicated collinearity. The final models’ performances were assessed through their discrimination capabilities using the Area Under the Receiver Operating Curve (AUC), which ranges from 0.5 (representing a discriminative ability equal to chance), to 1.0 (reflecting a perfect discriminative ability between those with and without the outcome). The evaluation of the models’ calibration, which determines the agreement between observed outcomes and predictive probabilities, was performed by analyzing calibration plots and slope. An ideal calibration is illustrated by a diagonal line on the graph with an intercept of 0 and a slope of 1. However, a slope less than 1 suggests overfitting in the model, meaning that respondents with high risk of the outcome have overestimated risk predictions while those with low risk of the outcome have underestimated risk predictions (18). Additionally, calibration-in-the-large (CITL) was assessed to identify the overall difference between the observed number of events and the average predictive risk.

LASSO for inference analysis was conducted, which allowed the reporting of ORs and CIs. A secondary analysis among Syrian migrants and refugees explored the association of regularized residency (possession of legal documents) on vaccination by logistic regression adjusted for confounders, including sex, education and years of living in Lebanon. All analyses were conducted using Stata/SE statistical software version 18 (STATACorp).

The study followed the TRIPOD reporting guidelines and STROBE reporting guidelines for prediction modelling (19, 20).

## Results

A total of 2,045 participants who completed the first wave of data collection were included in this analysis (Supp. Figure 1), of whom 79% were Lebanese (n=1,322), 18% Syrians (n=664) and 3% of other nationalities (n=59), while 0.8% (n=17) had a missing outcome. Median age of the study sample was 47 years (IQR 32-62, range 18-98) and 45.6% were males.

As of October 2022, COVID-19 vaccine uptake among study participants was 77%. There were differences in vaccination status between nationalities (Figure 1) with half (53%) of Syrians not having received any doses of the COVID-19 vaccine followed by other nationalities (39%) while 15% of Lebanese had not received any doses (P<0.001). In addition, the Lebanese population in this study had received a greater number of vaccine doses with 82% vaccinated having received 2 doses or more (vs. 39% and 55% for Syrians and other nationalities, respectively). Most common reasons reported for lack of vaccine uptake were: (1) not believing the vaccine is essential, and (2) preferring to follow other precautions - by both Lebanese (51% and 25%, respectively) and Syrians (45% and 23%, respectively). For those vaccinated with only one dose (3% of Lebanese and 8% of Syrians), the main reason was not wanting to receive the second dose for fear of side effects for Lebanese (44%), while among Syrian migrants and refugees the highest proportion stated they were still waiting for a second dose (41%) (Supp. Table 1).

**Figure 1.**
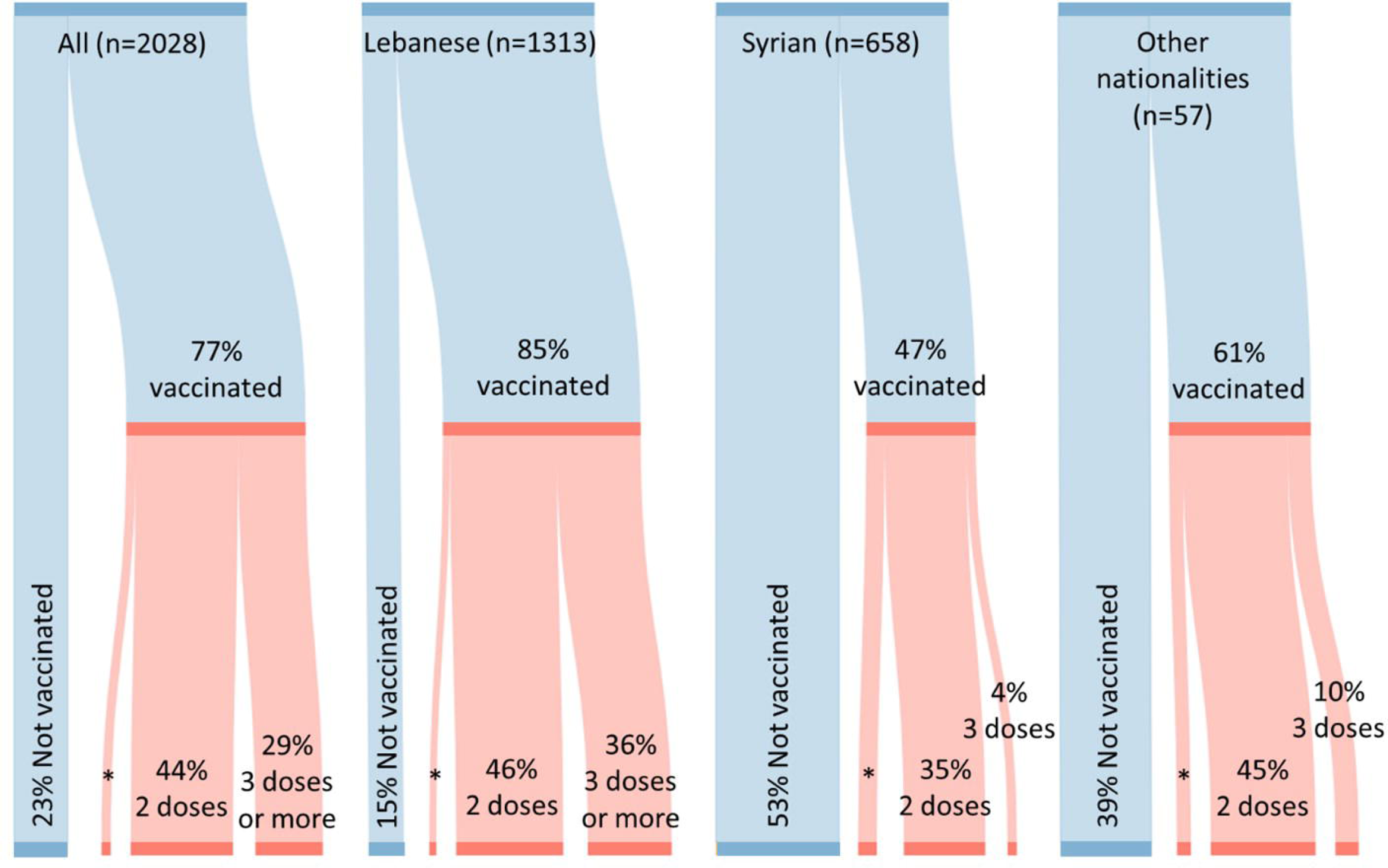
COVID-19 vaccine uptake among study participants. *\*Uptake of one dose of COVID-19 vaccine was 4% for the total sample, 3% for Lebanese, 8% for Syrians, and 6% for other nationalities*.

**Table 1.** Characteristics of the Lebanese population in our study area and associations with COVID-19 vaccine uptake.

|  | Total<br>(n=1,313) |  | Not vaccinated<br>(n=214) |  | Vaccinated<br>(n=1,099) |  | COVID-19<br>vaccine uptake |
| --- | --- | --- | --- | --- | --- | --- | --- |
|  | n | Weighted<br>% | n | Weighted<br>% | n | Weighted<br>% | Weighted<br>Unadjusted OR<br>[95% CI] |
| **Age (years) Median(IQR)** | 1304 | 52 (35-64) | 212 | 42 (33-57) | 1092 | 54 (36-65) | 1.02 [1.01-1.03] |
| Missing | 9 |  | 2 |  | 7 |  |  |
| **Sex** |  |  |  |  |  |  |  |
| Male | 588 | (45.0) | 93 | (43.8) | 495 | (45.2) | 1 |
| Female | 725 | (55.0) | 121 | (56.2) | 604 | (54.8) | 0.95 [0.70-1.28] |
| Missing | 0 |  | 0 |  | 0 |  |  |
| **Presence of an older adult in the household** |  |  |  |  |  |  |  |
| No | 565 | (40.3) | 125 | (55.7) | 440 | (37.5) | 1 |
| Yes | 743 | (59.7) | 89 | (44.3) | 654 | (62.5) | 2.10 [1.55-2.84] |
| Missing | 5 |  | 0 |  | 5 |  |  |
| **Education** |  |  |  |  |  |  |  |
| School not attended | 103 | (8.8) | 24 | (12.6) | 79 | (8.1) | 1 |
| School not completed | 578 | (47.6) | 89 | (43.8) | 489 | (48.3) | 1.72 [1.03-2.87] |
| School completed | 174 | (14.0) | 30 | (14.5) | 144 | (13.9) | 1.49 [0.81-2.75] |
| Vocational | 117 | (9.3) | 21 | (10.5) | 96 | (9.1) | 1.35 [0.70-2.61] |
| Higher Education | 251 | (20.3) | 38 | (18.6) | 213 | (20.6) | 1.73 [0.97-3.08] |
| Missing | 90 |  | 12 |  | 78 |  |  |
| **Assets-based wealth index** |  |  |  |  |  |  |  |
| 1 | 137 | (10.5) | 31 | (14.1) | 106 | (9.8) | 1 |
| 2 | 558 | (43.4) | 100 | (48.1) | 458 | (42.6) | 1.27 [0.80-2.03] |
| 3 | 587 | (46.1) | 77 | (37.8) | 510 | (47.6) | 1.81 [1.12-2.92] |
| Missing | 31 |  | 6 |  | 25 |  |  |
| **Presence of chronic illnesses** |  |  |  |  |  |  |  |
| No | 811 | (60.5) | 153 | (70.3) | 658 | (58.7) | 1 |
| Yes | 502 | (39.5) | 61 | (29.7) | 441 | (41.3) | 1.66 [1.20-2.30] |
| Missing | 0 |  | 0 |  | 0 |  |  |
| **Healthcare coverage** |  |  |  |  |  |  |  |
| Nothing | 669 | (53.8) | 145 | (69.3) | 524 | (50.9) | 1 |
| Private and mutual funds | 78 | (6.3) | 6 | (3.2) | 72 | (6.9) | 2.90 [1.23-6.84] |
| Public funds and NGOs/UN agencies | 487 | (39.9) | 55 | (27.5) | 432 | (42.2) | 2.08 [1.48-2.93] |
| Missing | 79 |  | 8 |  | 71 |  |  |
| **Believing COVID-19 is a serious infection** |  |  |  |  |  |  |  |
| True | 1121 | (86.6) | 168 | (79.9) | 953 | (87.8) | 1 |
| False | 172 | (13.4) | 41 | (20.1) | 131 | (12.2) | 0.55 [0.37-0.82] |
| Missing | 20 |  | 5 |  | 15 |  |  |
| **Consider themselves susceptible to a COVID-19 infection** |  |  |  |  |  |  |  |
| No | 310 | (23.5) | 75 | (34.9) | 235 | (21.4) | 1 |
| Yes | 997 | (76.5) | 136 | (65.1) | 861 | (78.6) | 1.97 [1.42-2.71] |
| Missing | 6 |  | 3 |  | 3 |  |  |
| **Vaccines are safe and/or effective** |  |  |  |  |  |  |  |
| Agree | 1023 | (78.3) | 86 | (39.0) | 937 | (85.4) | 1 |
| Neither agree or disagree | 182 | (13.7) | 56 | (26.8) | 126 | (11.4) | 0.19 [0.13-0.29] |
| Disagree | 106 | (8.0) | 71 | (34.2) | 35 | (3.2) | 0.04 [0.03-0.07] |
| Missing | 2 |  | 1 |  | 1 |  |  |
| **Previous receipt of the flu vaccine** |  |  |  |  |  |  |  |
| No | 915 | (73.7) | 169 | (83.4) | 746 | (71.9) | 1 |
| Yes | 328 | (26.3) | 34 | (16.6) | 294 | (28.1) | 1.96 [1.31-2.93] |
| Missing | 70 |  | 11 |  | 59 |  |  |

Data are n and weighted % or median (IQR), OR=odds ratio, CI=confidence interval.
Age was a continuous variable; increasing age was associated with increased odds of receiving the COVID-19 vaccine.

For Lebanese adults, older age, the presence of an older adult in the household, the highest tertile of asset-based wealth index, the presence of at least one multi-comorbidity, having private or public funds healthcare coverage, belief that COVID-19 is a serious infection, considering themselves susceptible to COVID-19, belief in the safety and efficacy of vaccines in general, and receipt of the flu vaccine were all associated with increased odds of receiving the COVID-19 vaccine (Table 1).

For Syrian migrants and refugees, similarly to Lebanese, older age, belief that COVID-19 is a serious infection, considering themselves susceptible to COVID-19, belief in the safety and efficacy of vaccines in general, and receipt of the flu vaccine were associated with increased odds of receiving the COVID-19 vaccine; while being male and having regularized residency were additional variables found to increase the odds of vaccination amongst Syrian migrants and refugees (Table 2). In a sub-group analysis amongst Syrian migrants and refugees, having legal residency status in Lebanon, a context specific factor to Syrian migrants and refugees, doubled the odds of COVID-19 vaccine uptake (adjusted OR:1.7 (95%CI:1.1-2.8)). An unadjusted bivariate analysis for other nationalities found COVID-19 vaccine uptake associated with attitudes towards COVID-19 and general vaccination (Supp. Table 2).

**Table 2.** Characteristics of the Syrian population in our study area and associations with COVID-19 vaccine uptake.

|  | **Total**<br>**(n=658)** |  | **Not vaccinated**<br>**(n=355)** |  | **Vaccinated**<br>**(n=303)** |  | **COVID-19**<br>**vaccine uptake** |
| --- | --- | --- | --- | --- | --- | --- | --- |
|  | **n** | **Weighted**<br>**%** | **n** | **Weighted**<br>**%** | **n** | **Weighted**<br>**%** | **Weighted**<br>**Unadjusted OR**<br>**[95% CI]** |
| **Age (years) Median(IQR)** | 656 | 35 (26-42) | 353 | 32 (24-41) | 303 | 37 (28-45) | 1.02 [1.01-1.04] |
| Missing | 2 |  | 2 |  | 0 |  |  |
| **Sex** |  |  |  |  |  |  |  |
| Male | 336 | (50.3) | 154 | (41.9) | 182 | (59.6) | 1 |
| Female | 322 | (49.7) | 201 | (58.1) | 121 | (40.4) | 0.49 [0.35-0.68] |
| Missing | 0 |  | 0 |  | 0 |  |  |
| **Education** |  |  |  |  |  |  |  |
| School not attended | 93 | (14.9) | 58 | (16.7) | 35 | (12.8) | 1 |
| School not completed | 450 | (71.2) | 248 | (72.5) | 202 | (69.7) | 1.25 [0.76-2.04] |
| School completed | 51 | (8.5) | 20 | (6.4) | 31 | (10.8) | 2.21 [1.03-4.72] |
| Vocational | 17 | (2.7) | 10 | (2.9) | 7 | (2.5) | 1.11 [0.36-3.42] |
| Higher Education | 17 | (2.7) | 6 | (1.5) | 11 | (4.2) | 3.45 [1.12-10.61] |
| Missing | 30 |  | 13 |  | 17 |  |  |
| **Receiving cash assistance** |  |  |  |  |  |  |  |
| No | 357 | (56.5) | 203 | (58.9) | 154 | (53.9) | 1 |
| Yes | 281 | (43.5) | 139 | (41.1) | 142 | (46.1) | 1.23 [0.88-1.71] |
| Missing | 20 |  | 13 |  | 7 |  |  |
| **Presence of chronic illness** |  |  |  |  |  |  |  |
| No | 533 | (79.8) | 289 | (79.3) | 244 | (80.4) | 1 |
| Yes | 125 | (20.2) | 66 | (20.7) | 59 | (19.6) | 0.93 [0.61-1.42] |
| Missing | 0 |  | 0 |  | 0 |  |  |
| **Healthcare coverage** |  |  |  |  |  |  |  |
| Nothing | 500 | (77.4) | 266 | (77.2) | 234 | (77.6) | 1 |
| Private, mutual and public funds | 3 | (0.4) | 2 | (0.5) | 1 | (0.3) | 0.56 [0.05-6.29] |
| NGOs/UN agencies | 143 | (22.2) | 79 | (22.3) | 64 | (22.1) | 0.98 [0.66-1.46] |
| Missing | 12 |  | 8 |  | 4 |  |  |
| **Believing COVID-19 is a serious infection** |  |  |  |  |  |  |  |
| True | 588 | (90.3) | 307 | (87.7) | 281 | (93.2) | 1 |
| False | 60 | (9.7) | 41 | (12.3) | 19 | (6.8) | 0.52 [0.28-0.97] |
| Missing | 10 |  | 7 |  | 3 |  |  |
| **Consider themselves susceptible to a COVID-19 infection** |  |  |  |  |  |  |  |
| No | 161 | (23.9) | 98 | (27.5) | 63 | (20.0) | 1 |
| Yes | 495 | (76.1) | 255 | (72.5) | 240 | (80.0) | 1.51 [1.03-2.23] |
| Missing | 2 |  | 2 |  | 0 |  |  |
| **Vaccines are safe and/or effective** |  |  |  |  |  |  |  |
| Agree | 444 | (68.4) | 183 | (51.8) | 261 | (86.8) | 1 |
| Neither agree or disagree | 142 | (20.8) | 106 | (29.0) | 36 | (11.8) | 0.24 [0.16-0.38] |
| Disagree | 69 | (10.8) | 64 | (19.2) | 5 | (1.4) | 0.04 [0.02-0.11] |
| Missing | 3 |  | 2 |  | 1 |  |  |
| **Previous receipt of the flu vaccine** |  |  |  |  |  |  |  |
| No | 449 | (72.5) | 260 | (78.4) | 189 | (65.9) | 1 |
| Yes | 184 | (27.5) | 80 | (21.6) | 104 | (34.1) | 1.88 [1.30-2.71] |
| Missing | 25 |  | 15 |  | 10 |  |  |
| **Have regularized residency** |  |  |  |  |  |  |  |
| No | 518 | (78.9) | 299 | (84.4) | 219 | (72.9) | 1 |
| Yes | 133 | (21.1) | 52 | (15.6) | 81 | (27.1) | 2.00 [1.31-3.05] |
| Missing | 7 |  | 4 |  | 3 |  |  |

*Data are n and weighted % or median (IQR), OR=odds ratio, CI=confidence interval.*
*Age was a continuous variable; increasing age was associated with increased odds of receiving the COVID-19 vaccine.*
*The chronic illnesses assessed included hypertension, type II diabetes, vascular diseases, dyslipidemia, chronic respiratory diseases, rheumatoid arthritis, chronic kidney diseases, cancer.*

The final models retained eight predictors of COVID-19 vaccine uptake for both the Lebanese and Syrian populations respectively. For Lebanese, identified predictors were older age, presence of an older adult in the household, higher education, greater asset-based wealth index, private healthcare coverage (i.e., having private or mutual insurance), feeling susceptible to COVID-19, belief in the safety and efficacy of vaccines, and previously received the flu vaccine (Table 3).

**Table 3.** Predictors of vaccine acceptance among Lebanese adults in a district of Beirut.

|  | Codes | Coefficients<sup>*</sup> | OR [95%CI]<sup>^</sup> |
| --- | --- | --- | --- |
| **Age (years)** |  | 0.02 | 1.03 [1.01-1.05] |
| **Presence of an older adult in the household** |  |  |  |
| No | 0 |  | Ref. |
| Yes | 1 | 0.46 | 1.73 [1.15-2.60] |
| **Education** |  |  |  |
| School not attended | 1 | -0.62 | Ref. |
| School not completed | 2 | - | 2.42 [1.16-5.05] |
| School completed | 3 | - | 2.51 [0.93-6.77] |
| Vocational | 4 | - | 2.67 [1.06-6.74] |
| Higher Education | 5 | 0.02 | 3.02 [1.26-7.21] |
| **Assets-based wealth index** |  |  |  |
| 1 | 1 | -0.08 | Ref. |
| 2 | 2 | - | 1.13 [0.64-1.99] |
| 3 | 3 | 0.46 | 1.97 [1.09-3.55] |
| **Healthcare coverage** |  |  |  |
| Nothing | 0 | -0.84 | Ref. |
| Private and mutual funds | 1 | 0.19 | 4.12 [1.40-12.13] |
| Public funds and NGOs/UN agencies | 2 | - | 2.55 [1.62-3.99] |
| **Consider themselves susceptible to a COVID-19 infection** |  |  |  |
| No | 0 |  | Ref. |
| Yes | 1 | 0.22 | 1.33 [0.87-2.02] |
| **Vaccines are safe and/or effective** |  |  |  |
| Agree | 1 | 1.57 | Ref. |
| Neither agree or disagree | 2 | - | 0.19 [0.12-0.30] |
| Disagree | 3 | -1.50 | 0.04 [0.02-0.07] |
| **Previous receipt of the flu vaccine** |  |  |  |
| No | 0 |  | Ref. |
| Yes | 1 | 0.04 | 1.17 [0.60-1.96] |
| **Intercept** |  | -0.25 |  |
| **AUC** |  |  | 0.823 [0.786-0.859] |
| **C-Slope** |  |  | 1.061 [0.905-1.217] |
| **CITL** |  |  | 0.004 |

*AUC=Area Under the Receiver Operating Curve, C-Slope=Calibration-slope, CITL=calibration-in-the-large.*
*OR=odds ratio, CI=confidence interval.*
*\*Obtained from the LASSO logistic regression analysis. ^Obtained from the LASSO for inference analysis.*

While among Syrian migrants and refugees, the predictors were greater age, male sex, having completed school or higher education, receipt of cash assistance, presence of co-morbidities, belief in the safety and efficacy of vaccines, previously received the flu vaccine, and legal residency status in Lebanon (Table 4).

**Table 4.** Predictors of vaccine acceptance among Syrian adults in a district of Beirut.

|  | Codes | Coefficients* | OR [95%CI]^ |
| --- | --- | --- | --- |
| **Age (years)** |  | 0.02 | 1.03 [1.01-1.05] |
| **Sex** |  |  |  |
| Male | 0 | - | Ref. |
| Female | 1 | -0.51 | 0.51 [0.34-0.74] |
| **Education** |  |  |  |
| School not attended | 1 | -0.03 | Ref. |
| School not completed | 2 | - | 1.25 [0.68-2.30] |
| School completed | 3 | 0.43 | 2.49 [1.00-6.16] |
| Vocational | 4 | - | 1.25 [0.38-4.16] |
| Higher Education | 5 | 0.39 | 3.02 [0.64-14.29] |
| **Receiving cash assistance** |  |  |  |
| No | 0 | - | Ref. |
| Yes | 1 | 0.19 | 1.52 [1.00-2.30] |
| **Presence of chronic illness** |  |  |  |
| No | 0 | - | Ref. |
| Yes | 1 | 0.02 | 1.21 [0.73-2.02] |
| **Vaccines are safe and/or effective** |  |  |  |
| Agree | 1 | 1.26 | Ref. |
| Neither agree or disagree | 2 | - | 0.25 [0.15-0.41] |
| Disagree | 3 | -1.12 | 0.05 [0.01-0.14] |
| **Previous receipt of flu vaccine** |  |  |  |
| No | 0 | - | Ref. |
| Yes | 1 | 0.35 | 1.68 [1.10-2.56] |
| **Have regularized residency** |  |  |  |
| No | 0 | - | Ref. |
| Yes | 1 | 0.51 | 2.07 [1.21-3.56] |
| **Intercept** |  | -1.77 |  |
| **AUC** |  |  | 0.768 [0.730-0.806] |
| **C-Slope** |  |  | 1.199 [0.955-1.442] |
| **CITL** |  |  | -0.012 |

*AUC=Area Under the Receiver Operating Curve, C-Slope=Calibration-slope, CITL=calibration-in-the-large.*
*OR=odds ratio, CI=confidence interval.*
*\*Obtained from the LASSO logistic regression analysis. ^Obtained from the LASSO for inference analysis.*

Both models showed good discrimination with an AUC of 0.823 (95%CI:0.786-0.859) for the Lebanese sample and 0.768 (95%CI:0.730-0.806) for the Syrian sample. The model for the Lebanese population had good calibration with a calibration slope of 1.061 (95%CI:0.905-1.217) and CITL of 0.004 (Supp. Figure 2). The model for the Syrian population had underfitting with a calibration slope of 1.199 (95%CI:0.955-1.442) and CITL of −0.012 (Supp. Figure 3).

To illustrate the model amongst Syrian migrants and refugees, a 40-year-old male Syrian with no legal documentation, who completed school, is not receiving cash assistance, does not have a co-morbidity, does not believe in the safety and efficacy of vaccines, and has never taken the flu vaccine has a 4% chance of having taken the COVID-19 vaccine.

## Discussion

This study revealed inequalities in COVID-19 vaccination status between Syrian migrants and refugees and Lebanese adults residing in a low socioeconomic area in Beirut. Overall, 53% of Syrian migrant and refugee adults had not received any doses of the COVID-19 vaccine compared to 15% among Lebanese adults. This study also developed and internally validated a prediction model that identified eight predictors of COVID-19 vaccine uptake of at least one dose for the Lebanese and Syrian populations, respectively. This study found socioeconomic, demographic and health risk perceptions to predict vaccine uptake for both Syrians and Lebanese. In addition, legal residency status in the country and sex were found to lead to differentials in vaccine uptake within the Syrian population.

The proportion of vaccinated Syrian migrants and refugees in this sample was similar to another study examining COVID-19 vaccination in Syrian refugees aged 50 years or older in Lebanon, which reported 42.5% having received at least one dose of the COVID-19 vaccine by mid-March 2022 (5). Furthermore, vaccination status in Syrian migrants and refugees in this sample was slightly lower than the 50% national COVID-19 vaccination average recorded at the time of data collection (6). In contrast, the proportion of vaccinated Lebanese reported in this study at 85% exceeds this national prevalence, and the majority (82%) were vaccinated with 2 doses or more which reached the national immunization target of 70% (3). These findings demonstrate large differentials in vaccination uptake by nationality, which indicate barriers to vaccination uptake amongst Syrian migrants and refugees. Thus, understanding the reasons for not taking the vaccine remains important to design contextualized public health interventions.

A previous study conducted on older Syrian refugees living in Lebanon found similar predictors, bar the direction of the association of age and vaccine uptake; likely due to the larger age range included in the present study (5). To the best of our knowledge, there are no prediction models for COVID-19 vaccine uptake among Lebanese adults or younger Syrian migrants and refugees.

Some of the identified predictors of COVID-19 vaccine uptake in this study, older age and education level, were common for both Lebanese (national) and Syrian (non-national) populations, which was consistent with studies conducted in the MENA region (21, 22), the UK (10), the US (11, 23, 24), and a global meta-analysis (25). In general, older adults have physiological vulnerabilities and co-morbidities, and thus may perceive themselves at higher risk of infection and severe complications (21, 23, 25). Higher education level was associated with higher likelihood of vaccine uptake (5) and higher willingness to receive the vaccine (10, 21–26). This may be explained by those with a higher educational level being more aware of the risks of the disease, the importance of vaccination and having the ability to critically appraise vaccine misinformation (12, 23). Health and COVID-19 risk beliefs and perceptions, as well as trust in vaccination effectiveness following positive past experiences, also played a role in vaccine acceptance and uptake in line with other studies (11, 21, 23–25, 27).

The socio-economic indicators identified, where both Lebanese in the highest tertile of wealth and Syrian migrants and refugees receiving cash assistance had a higher likelihood of vaccine uptake, were in accordance with studies in low and middle-income countries reported low socio-economic status as a determinant of low willingness to take the COVID-19 vaccine (26). Studies in other context also align, as in the US low income was associated with lower vaccine uptake and greater vaccine hesitancy (23, 24). Despite the COVID-19 vaccine being available free of charge to all residents (3), this highlights that there may be other barriers to attaining the vaccine specifically for Syrian migrants and refugees.

Legal residency status (having a legal residency permit in Lebanon) was found as a context specific predictor of COVID-19 vaccine uptake for Syrian migrants and refugees. The majority of the Syrian migrants and refugees (79%) reported not having a legal residency permit; this was likely due to the high costs and requirements relating to obtaining and renewing annual residency permits (8). Previous studies in other countries have also shown that lack of legal residency permit among migrants impacts health care access (28). Consequently, lack of legal residency may have impacted vaccination uptake as residents of any nationality in Lebanon were required to register on an online government platform (3), thus Syrian migrants and refugees, particularly those without documentation, may have worried that registering may have led to arrest or deportation (8, 28, 29).

### Strengths and Limitations

This study has several strengths and limitations. The study outcome was self-reported which may be prone to information bias; however, recall of vaccinations amongst adults was shown to be relatively accurate (30). The study sample may not be representative of all adults in Lebanon, yet it is representative of this low socio-economic suburb in Beirut. The model was slightly underfitted for Syrians and a larger sample may help prevent this in the future, whilst this study had a large enough sample size of Lebanese adults to be able to develop a well calibrated model.

### Conclusion

By October 2022, more than half the Syrian migrants and refugees living in a low socio-economic area of a suburb of Beirut were still not vaccinated against COVID-19 despite the availability of vaccines free of charge to all residents including non-nationals in Lebanon, as opposed to the majority of Lebanese from the same area having received the vaccine. Predictors for both populations include older age, educational level, socio-economic status, and general attitude toward vaccination. The findings indicate barriers for vaccine uptake in Syrian migrants and refugees, including legal status. This calls for urgent action to eliminate barriers and enable equitable access to vaccines for all vulnerable population in outbreaks such as the COVID-19 pandemic.

## Contributors

HG, JD and SJM conceptualized the study; SJM, HG and MER designed the survey, contribution to study methodology and investigation, and oversaw the data collection. The formal analysis and literature search was conducted by MER. MER and SJM drafted the manuscript. SJM supervised MER throughout the project. MER, HG, HS, JD, NM and SJM contributed to the interpretation of the results, and have edited drafts and approved the final version of the Article. MER, HG and SJM had full access to and verified the raw data. All authors had access to the study data and had final responsibility for the decision to submit the manuscript for publication.

## Data sharing

The anonymized data can be obtained upon reasonable request from the Center for Research on Population and Health at the American University of Beirut.

## Acknowledgments

We would like to thank the “Community Action for Equity in Pandemic preparedness and control” CAEP Study Group for supporting this work (Aline Germani, Fadi El-Jardali). We thank Berthe Abi Zeid for her support and expertise in data analysis. We acknowledge BOT (Bridge. Outsource. Transform) for their assistance in collecting the data required for the success of this study and the study participants for their participation.

