## Supplementary material for "Development of Prediction Models of COVID-19 Vaccine Uptake among Lebanese and Syrians in a district of Beirut, Lebanon: a population-based study"

**Supp. Table 1 – Reasons for COVID-19 vaccine uptake hesitancy in our study sample across nationalities**

|  | **Total** | | **Lebanese** | | **Syrian** | | **Other Nationalities** | | **P-value** |
| --- | --- | --- | --- | --- | --- | --- | --- | --- | --- |
|  | *n* | *Weighted %* | *n* | *Weighted %* | *n* | *Weighted %* | *n* | *Weighted %* |  |
| **For those unvaccinated, the main reason why they did not receive the COVID-19 vaccine** |  |  |  |  |  |  |  |  |  |
| Plan to register on platform | 56 | (8.5) | 10 | (4.5) | 43 | (13.2) | 3 | (15.7) | 0.013 |
| Prefer other precautionary measures | 121 | (23.0) | 50 | (24.9) | 71 | (23.5) | 0 | (0.0) |  |
| Do not believe vaccine is essential | 252 | (49.3) | 103 | (51.5) | 137 | (44.7) | 12 | (59.3) |  |
| Do not believe COVID-19 requires a vaccine | 46 | (9.2) | 17 | (8.3) | 25 | (9.0) | 4 | (20.5) |  |
| Other* | 52 | (10.0) | 21 | (10.8) | 30 | (9.6) | 1 | (4.5) |  |
| **Reasons only one dose of the COVID-19 vaccine was taken** |  |  |  |  |  |  |  |  |  |
| Medical condition/Doctor advise | 9 | (7.6) | 3 | (6.4) | 6 | (10.7) | 0 | (0.0) | 0.129 |
| Not wanting to for fear of side effects | 31 | (35.3) | 17 | (43.9) | 14 | (24.8) | 0 | (0.0) |  |
| Think one dose is enough | 25 | (27.5) | 13 | (29.0) | 10 | (19.7) | 2 | (67.1) |  |
| Waiting for second dose | 27 | (28.3) | 8 | (20.7) | 18 | (41.1) | 1 | (32.9) |  |
| Other | 2 | (1.3) | 0 | (0.0) | 2 | (3.7) | 0 | (0.0) |  |

*A P-value less than 0·05 was considered statistically significant.*

**Other reasons include: Mistrust, fear or hesitancy because of controversial information and the vaccine being new ; Medical reasons*

**Supp. Table 2 - Characteristics of other nationalities in our study area and associations with COVID-19 vaccine uptake**

|  | **Total**  **(n=57)** | | **Not vaccinated**  **(n=22)** | | **Vaccinated**  **(n=35)** | | **COVID-19**  **vaccine uptake** |
| --- | --- | --- | --- | --- | --- | --- | --- |
|  | **n** | **Weighted**  **%** | **n** | **Weighted**  **%** | **n** | **Weighted**  **%** | **Weighted**  **Unadjusted OR [95% CI]** |
| **Age (years)** **Median**(IQR) | 57 | 35 (30-46) | 22 | 31 (27-35) | 35 | 40 (34-52) | 1.06 [0.97-1.16] |
| **Sex** |  |  |  |  |  |  |  |
| Male | 18 | (33.0) | 9 | (42.9) | 9 | (26.5) | 1 |
| Female | 39 | (67.0) | 13 | (57.1) | 26 | (73.5) | 2.08 [0.63-6.89] |
| Missing | 0 |  | 0 |  | 0 |  |  |
| **Presence of an older adult in the household** |  |  |  |  |  |  |  |
| No | 47 | (80.3) | 18 | (79.3) | 29 | (81.0) | 1 |
| Yes | 10 | (19.7) | 4 | (20.7) | 6 | (19.0) | 0.90 [0.21-3.85] |
| Missing | 0 |  | 0 |  | 0 |  |  |
| **Education** |  |  |  |  |  |  |  |
| School not attended | 18 | (33.5) | 6 | (29.6) | 12 | (36.2) | 1 |
| School not completed | 21 | (43.5) | 11 | (54.4) | 10 | (35.8) | 0.54 [0.14-2.12] |
| School completed | 3 | (6.3) | 1 | (5.2) | 2 | (7.2) | 1.14 [0.08-16.88] |
| Vocational | 1 | (2.4) | 0 | (0.0) | 1 | (4.1) | . |
| Higher Education | 7 | (14.3) | 2 | (10.8) | 5 | (16.7) | 1.27 [0.17-9.43] |
| Missing | 7 |  | 2 |  | 5 |  |  |
| **Assets-based wealth index** |  |  |  |  |  |  |  |
| 1 | 37 | (66.2) | 13 | (62.6) | 24 | (68.5) | 1 |
| 2 | 11 | (19.3) | 5 | (22.8) | 6 | (17.1) | 0.69 [0.16-2.93] |
| 3 | 8 | (14.5) | 3 | (14.6) | 5 | (14.4) | 0.90 [0.17-4.70] |
| Missing | 1 |  | 1 |  | 0 |  |  |
| **Receiving cash assistance** |  |  |  |  |  |  |  |
| No | 49 | (87.7) | 20 | (95.0) | 29 | (83.1) | 1 |
| Yes | 7 | (12.3) | 1 | (5.0) | 6 | (16.9) | 3.86 [0.40-37.56] |
| Missing | 1 |  | 1 |  | 0 |  |  |
| **Presence of chronic illness** |  |  |  |  |  |  |  |
| No | 47 | (81.6) | 19 | (84.9) | 28 | (79.5) | 1 |
| Yes | 10 | (18.4) | 3 | (15.1) | 7 | (20.5) | 1.45 [0.31-6.75] |
| Missing | 0 |  | 0 |  | 0 |  |  |
| **Believing COVID-19 is a serious infection** |  |  |  |  |  |  |  |
| True | 45 | (82.7) | 15 | (71.9) | 30 | (89.3) | 1 |
| False | 10 | (17.3) | 6 | (28.1) | 4 | (10.7) | 0.31 [0.07-1.38] |
| Missing | 2 |  | 1 |  | 1 |  |  |
| **Consider themselves susceptible to a COVID-19 infection** |  |  |  |  |  |  |  |
| No | 22 | (39.9) | 12 | (57.7) | 10 | (28.4) | 1 |
| Yes | 35 | (60.1) | 10 | (42.3) | 25 | (71.6) | 3.43 [1.06-11.07] |
| Missing | 0 |  | 0 |  | 0 |  |  |
| **Vaccines are safe and/or effective** |  |  |  |  |  |  |  |
| Agree | 37 | (68.2) | 7 | (35.8) | 30 | (86.2) | 1 |
| Neither agree or disagree | 8 | (13.9) | 5 | (25.3) | 3 | (7.6) | 0.12 [0.02-0.74] |
| Disagree | 9 | (17.9) | 7 | (38.9) | 2 | (6.2) | 0.07 [0.01-0.42] |
| Missing | 3 |  | 3 |  | 0 |  |  |
| **Previous receipt of the flu vaccine** |  |  |  |  |  |  |  |
| No | 39 | (79.3) | 18 | (97.7) | 21 | (68.0) | 1 |
| Yes | 12 | (20.7) | 1 | (2.3) | 11 | (32.0) | 20.02 [2.16-185.73] |
| Missing | 6 |  | 3 |  | 3 |  |  |

*Data are n and weighted % or median (IQR), OR=odds ratio. CI=confidence interval.*

*Age was a continuous variable.*

*The wealth index was generated from the presence in the household of functional transportation, communication, home technology and cooking assets.*

*The chronic illnesses assessed included hypertension, type II diabetes, vascular diseases, dyslipidemia, chronic respiratory diseases, rheumatoid arthritis, chronic kidney diseases, cancer.*

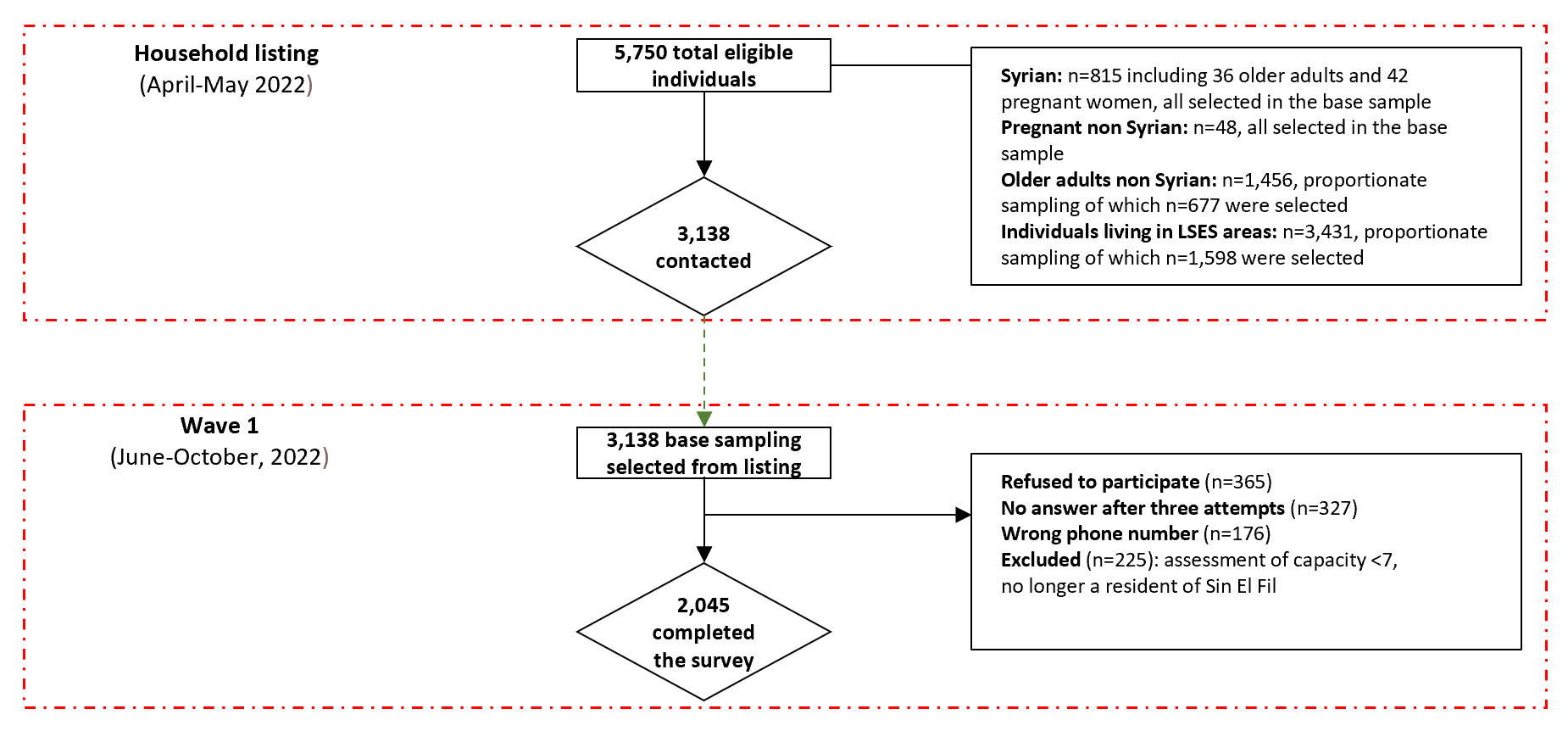

**Supp. Figure 1. Flow diagram of the study population who completed the survey at wave 1**

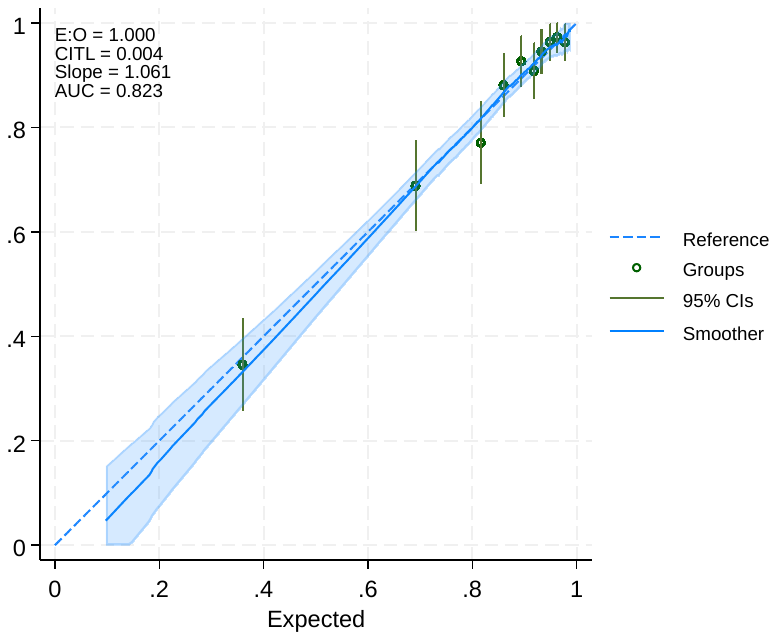

**Supp. Figure 2. Final model performance for the Lebanese sample**

*E:O= Expected to Observed ratio (calibration) ; CITL= calibration-in-the-large ; Slope=Calibration-slope ; AUC= Area Under the Curve (discrimination)*

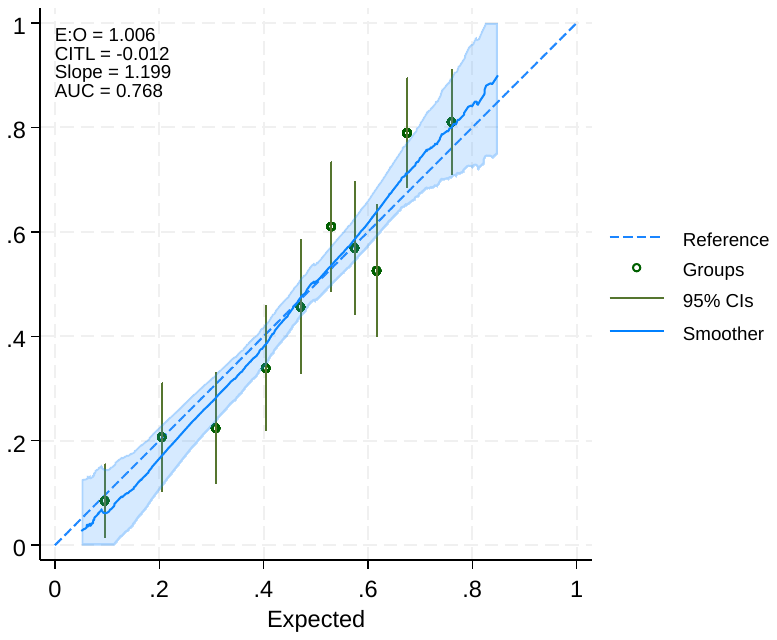

**Supp. Figure 3. Final model performance for the Syrian sample**

*E:O= Expected to Observed ratio (calibration) ; CITL= calibration-in-the-large ; Slope=Calibration-slope ; AUC= Area Under the Curve (discrimination)*
